# Influence of menstrual cycle and oral contraceptive phases on bone (re)modelling markers in response to intervallic running

**DOI:** 10.1101/2023.11.29.23299199

**Authors:** Isabel Guisado-Cuadrado, Nuria Romero-Parra, Kirsty J. Elliott-Sale, Craig Sale, Ángel E. Díaz, Ana B. Peinado

## Abstract

**Objectives:** To examine procollagen type I N-propeptide (P1NP) and carboxy-terminal cross-linking telopeptide of type I collagen (β-CTX-1) concentrations across different phases of the menstrual (MC) and oral contraceptive (OC) cycles and in response to running.

**Methods:** 17β-oestradiol, progesterone, P1NP and β-CTX-1 were analysed pre-and post-exercise in eight eumenorrheic females in the early-follicular (EFP), late-follicular (LFP), and mid-luteal (MLP) phases of the MC, while 8 OC users were evaluated during the withdrawal (WP) and active pill-taking (APP) phases. The running protocol consisted of 8x3 min treadmill runs at 85% of maximal aerobic speed.

**Results:** 17β-oestradiol concentrations (pg·ml^-1^) were lower in the EFP (47.22±39.75) compared to the LFP (304.95±235.85; p=<0.001) and MLP (165.56±80.6; p=0.003) of the MC and higher in the WP (46.51±44.09) compared to the APP (10.88±11.24; p<0.001) of the OC cycle. While progesterone (ng·ml^-1^) was higher in the MLP (13.214±4.926) compared to the EFP (0.521±0.365; p<0.001) and LFP (1.677±2.586; p<0.001) of the MC. In eumenorrheic females, P1NP concentrations (ng·ml^-1^) were higher in LFP (69.97±17.84) compared to EFP (60.96±16.64; p=0.006;) and MLP (59.122±11.77; p=0.002;). Post-exercise concentrations (70.71±15.59) increased from pre-exercise (55.86±12.86; p<0.001). For β-CTX-1 (ng·ml^-1^), lower concentrations were shown in MLP (0.376±0.098) compared to LFP (0.496±0.166; p=0.001) and EFP (0.452±0.148; p=0.039). OC users showed higher post-exercise P1NP concentrations in WP (61.75±8.32) compared to post-exercise in APP (45.45±6; p<0.001). Comparing P1NP levels between eumenorrheic and OC users, post-exercise P1NP concentrations were higher in the EFP (66.91±16.26; p<0.001), LFP (80.66±16.35; p<0.001) and MLP (64.57±9.68; p=0.002) of the MC compared to the APP of the OC cycle.

**Conclusion:** These findings underscore the importance of studying exercising females with different ovarian hormone profiles, as changes in sex hormone concentrations affect bone metabolism in response to running, showing a higher post-exercise P1NP concentrations in all MC phases compared with APP of the OC cycle.

## 1. Introduction

Exercise is commonly considered beneficial for bone health and a preventive strategy for age-related bone loss^1^ or to achieve a higher peak bone mass during the growth stage.^2^ Exercise characteristics are, however, important to produce an osteogenic stimulus, including multidirectional movements, high impact intensity and load changes during training.^3,4^ On the contrary, participation in endurance sports that involve repetitive or lower impact loads (e.g., running) or non-weight bearing sports (e.g., cycling or swimming) generally do not produce skeletal benefits,^5^ although some controversy remains about the osteogenic effect of running.^6^ Furthermore, bone stress injuries are a concern that directly affects female athletes, as their incidence is higher than in men (for a review, please see^7^). These injuries occur when excessive repetitive loads are introduced into bone tissue and an imbalance in bone metabolism favours the accumulation of microdamage over its removal and replacement by new bone tissue during the remodelling process (for a review, please see^7^).

Beyond its reproductive function, 17β-oestradiol has an important effect on bone cells, producing a longer osteoblast lifespan and an increase in their metabolic activity, thus participating in the bone formation/resorption balance.^8^ Consequently, menstrual status is directly linked to bone health; menstrual disturbances have been identified as a risk factor for stress-related bone injuries and lower bone mineral density (BMD).^9,10^ Some studies suggest that hormonal fluctuations throughout the menstrual cycle (MC) should be considered when measuring bone remodelling markers,^11^ although not all studies agree.^12,13^

Given the high prevalence of OC use among female athletes,^14^ different hormonal profiles should be considered when assessing bone health. During the hormonal active pill-taking phase (APP) 17α-ethinyl oestradiol inhibits endogenous 17β-oestradiol production, while in the placebo or withdrawal phase (WP), endogenous 17β-oestradiol increases again.^15^ Nevertheless, there is limited evidence relating to the effect of taking OCs and the associated implications (exogenous hormone supply and reduction of endogenous sex hormones) on bone (re)modelling markers, as well as their influence on the post-exercise bone remodelling markers concentrations.

A major challenge in assessing the bone metabolic response to a specific exercise intervention has been the methodology, since both BMD assessment, by dual-energy X-ray absorptiometry (DXA), and microarchitectural assessment, by computed tomography, require long-term interventions before changes become apparent, ^16^ which increases the potential for many confounding factors to exert an affect. Thus, bone (re)modelling marker measurements have been used as an alternative to assess acute responses to a stimulus, such as exercise, although some questions remain over how they relate to longer-term outcomes (BMD, material and structural properties, etc.).^17,18^ Specifically, the International Osteoporosis Foundation and the International Federation of Clinical Chemistry and Laboratory Medicine recommend the use of procollagen type I N-propeptide (P1NP) and carboxy-terminal cross-linking telopeptide of type I collagen (β-CTX-1) concentrations, as reference markers of bone formation and bone resorption.^11^ Therefore, this study aimed to examine the P1NP and β-CTX-1 responses to high-intensity running in eumenorrheic females and OC users.

## 2. Material and methods

### 2.1. Participants

Participants included in this study were part of the IronFEMME project, which received ethical clearance from the Research Ethics Committee of the Universidad Politécnica de Madrid. The purpose of IronFEMME was to determine the influence of sex hormones on iron metabolism and muscle damage, hence, the present study is a secondary analysis that was carried out after the trial was completed. This trial was registered at clinicaltrials.gov (ID: NCT04458662). To be included in the IronFEMME study, participants were required to meet the following criteria: (i) healthy adult females between 18 and 40 years; (ii) regular MCs (defined as normally occurring MCs from 21 to 35 days in length)^19^ at least 6 months prior to the study ; (iii) or using monophasic combined OC pills for at least 6 months prior to the study; (iv) no regular consumption of medication or nutritional supplements; (v) non-smokers; (vi) non-pregnant or oophorectomized; and (vii) participating in endurance training between 3 and 12 h per week. By using blood samples collected as part of the IronFEMME study, the present trial was designed as a secondary analysis, for which the inclusion criteria were further narrowed beyond those determined for the IronFEMME project. These additional criteria were (i) age between 20 and 32 years; (ii) not taking collagen supplements, calcium, or any substance that interferes/participates in bone metabolism; (iii) not having suffered any bone fracture for at least one year prior to the start of the study; and (iv) participating in endurance training involving running (i.e., long distance running, trail running, triathlon) between 3 and 12 h per week (see Table 1 for training volume). Therefore, the study sample was limited to eight eumenorrheic females and eight monophasic OC users (see Table 1 for participants’ characteristics and training volume). All participants were informed of the study procedures (*i.e.*, for the present study on bone (re)modelling) and risks prior to participation and written informed consent was obtained from each subject prior to inclusion. Participants also agreed to the use of their data for other scientific purposes *a posteriori*, which applies to the present study.

**Table 1.**
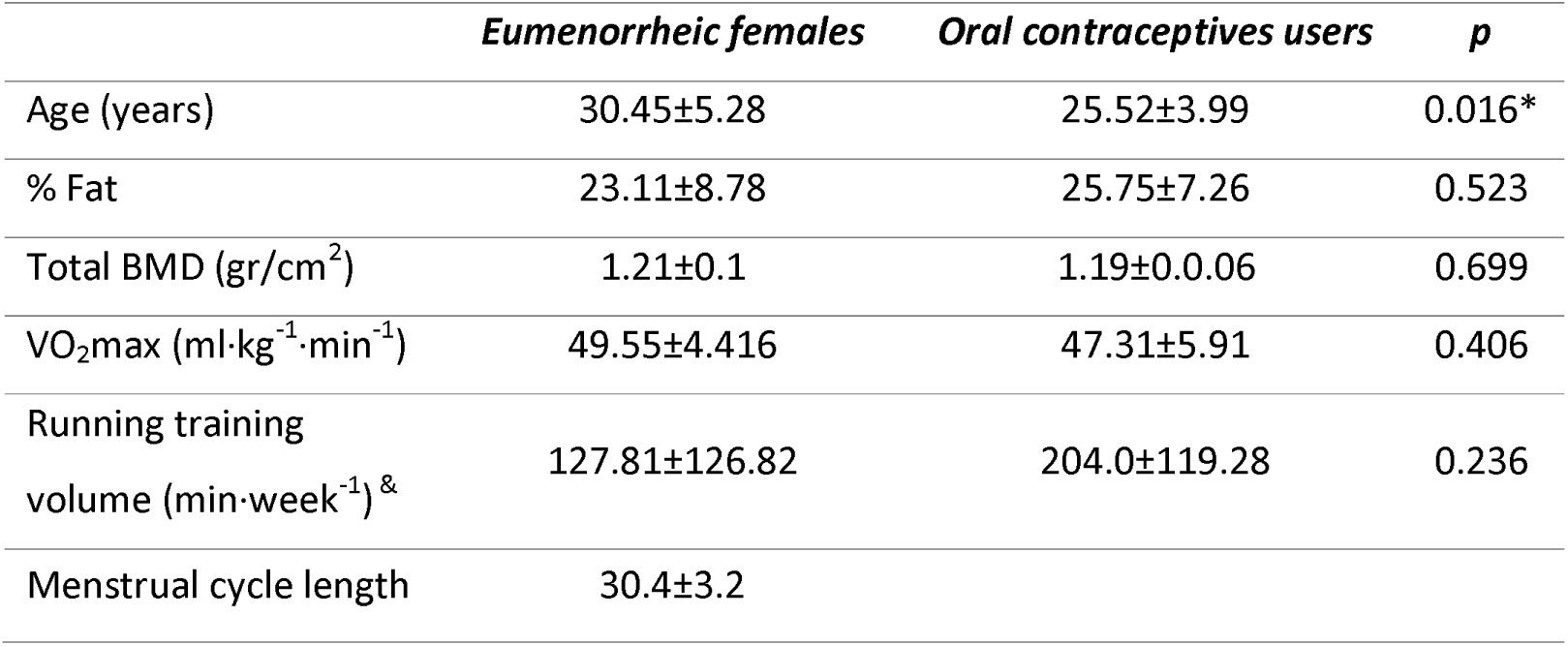
Participant characteristics show as mean±SD. ^&^Training volume during the 6 months prior to recruitment. Eumenorrheic participants = 8, Oral contraceptive participants = 8. VO2max = maximal oxygen uptake; BMD = total bone mineral density. * Significant differences between groups (p<0.05).

### 2.2. MC and OC cycle monitoring

MC monitoring was based on the three-step methodology. The theoretical MC phases were predicted by a gynecologist using the calendar-based counting method, based on records of the length of each participant’s last six MCs. Secondly, a urine-based predictor kit (Ellatest, Alicante, Spain) was used to identify the LH surge and subsequent ovulation. Participants collected their mid-morning urine (always at the same time of day) starting three to five days before the estimated LFP testing day until the test result was positive. A participant was excluded from the trial if a positive LH test result was not obtained in three MCs, was as they were considered to have anovulatory MCs, and if the progesterone concentration in the MLP was lower than 16 nmol/L. Finally, all phases were confirmed by serum sex hormone analysis taken on study days prior to the exercise bout. The EFP was characterised by lower levels of 17β-oestradiol and progesterone. The LFP was characterised by higher 17β-oestradiol concentrations than in the EFP and MLP and higher progesterone concentrations than in the EFP, but lower than 6.36 nmol/L. The MLP was characterised by a progesterone concentration greater than 16 nmol/L.

OC users took their active hormone pill daily for 21 days during the active pill-taking phase, followed by a 7-day withdrawal phase (pill without hormonal content). The mean duration of the OC use was 4.09±2.93 years (mean±SD). The brands and dosages of exogenous sex hormones in the monophasic combined OC preparations used by these participants were as follows: Yasmin® (n=2): 0.03 mg ethinyl oestradiol and 3 mg drospirenone; Linelle® (n=2): 0.02 mg ethinyl oestradiol and 0.1 mg levonorgestrel; Sibilla® (n=2): 0.03 mg ethinyl oestradiol and 2 mg dienogest (n=2); Edelsin® (n=1): 0.035 mg ethinyl oestradiol and 25 mg norgestimate; and Yasminelle® (n=1): 0.02 mg ethinyl oestradiol and 3 mg drospirenone.

### 2.3. Experimental overview

Eumenorrheic participants came to the laboratory on four occasions (Figure 1), the first for a maximal incremental treadmill test and the following three times to perform the intervallic running test in each phase of the MC phases (EFP, LFP and MLP). The EFP testing session took place on day 4±1 of the MC. The LPF testing session took place 2 days prior to predicted ovulation, on the day 12±2 of the MC; predicted ovulation was based on previous cycles in which ovulation was confirmed. If ovulation did not occur within 2 days of the predicted LPF testing session, the testing session was deemed invalid. The MLP took place on the day 21±3 of the MC. OC users came to the laboratory on 3 occasions, the first for the maximal incremental test and the following 2 times to carry out the intervallic test in the WP (day 6±1) and APP (day 22±5) of the OC cycle. The first session consisted of participants screening, while on the following sessions participants performed the interval running test in each of the MC and OC cycle phases in a randomized and counterbalanced manner. In the eumenorrheic group, the order of performance of the intervallic tests was randomized according to the phases of the MC as follows: EFP-LFP-MLP; LFP-MLP-EFP; MLP-EFP-LFP; LFP-EFP-MLP and EFP-MLP-LFP. For the group of OC users, the randomization was: WP-APP and APP-WP.

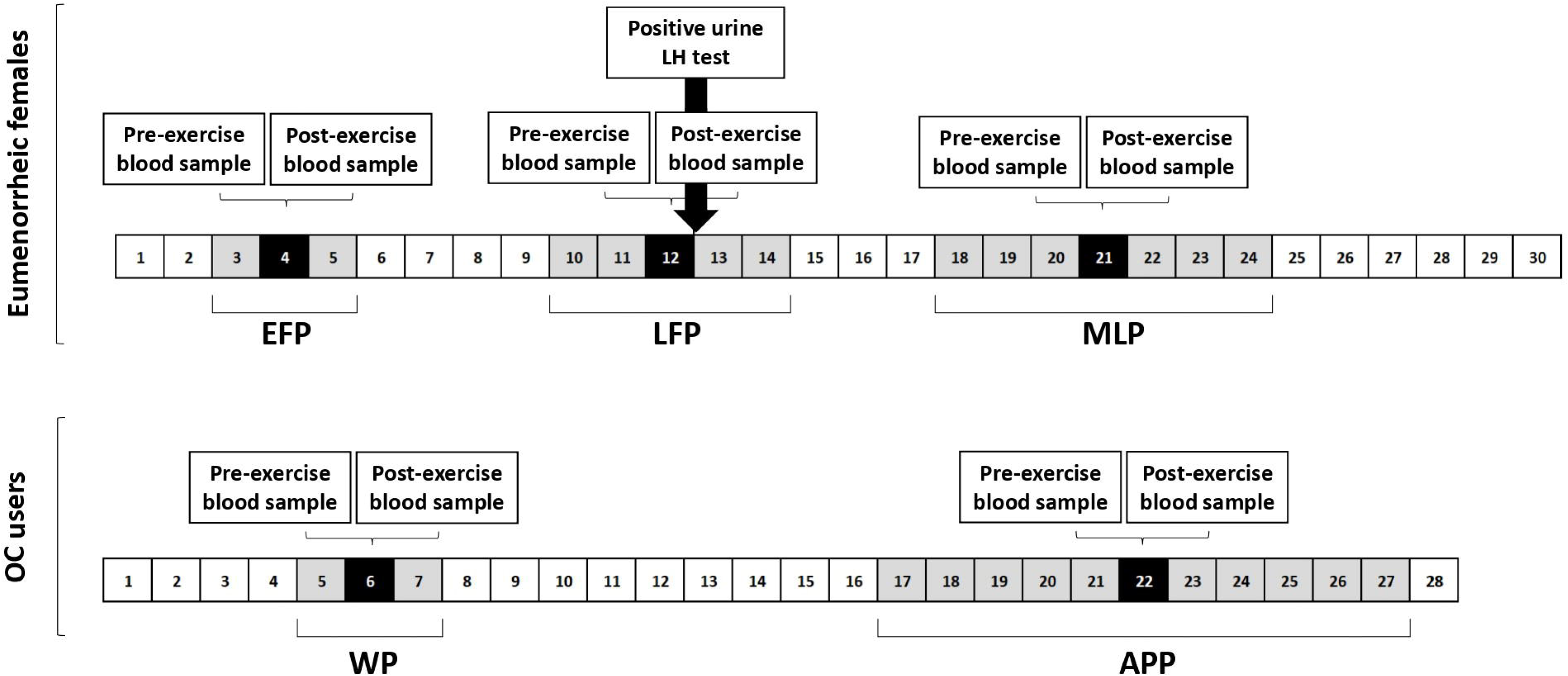

On day of screening, volunteers attended the laboratory between 8:00 a.m. and 10:00 a.m. in a resting and fasting state: during the EFP in the eumenorrheic group and day 4-7 of the WP in the OC users. Baseline antecubital venous blood samples were collected for complete blood count, biochemical, and hormonal analysis. After collecting the blood sample, a total body DXA was performed. This screening session was completed with an incremental running exercise to exhaustion on a computerised treadmill (H/P/COSMOS 3PW 4.0, H/P/Cosmos Sports & Medical, Nussdorf-Traunstein, Germany) to determine their maximal oxygen uptake. Expired gases were measured breath-by-breath with a Jaeger Oxycon Pro gas analyser (Erich Jaeger, Viasys Healthcare, Friedberg, Germany). This incremental maximal protocol began with a 3 min warm-up at 6 km/h followed by the incremental test in which the initial speed was set at 8 km/h and then increased by 0.2 km/h every 12 s until exhaustion. Prior to the maximal aerobic test and all the intervallic running tests all participants were instructed to refrain from alcohol, caffeine, and any intense physical activity or sport 24 hours before to visit the laboratory.

### 2.4. Intervallic running protocol

After the screening day in which the maximal incremental treadmill test was performed with the objective of determining maximal aerobic speed (vVO2peak), interval running tests were performed based upon the obtained values. This intervallic protocol consisted of a 5-min warm-up at 60% of the vVO2peak followed by eight bouts of 3 min at 85% of the vVO2peak with 90-s recovery at 30% of the vVO2peak between bouts. Finally, a 5-min cooling down was performed at 30% of vVO2peak. The intervallic tests were performed in a maximum of two consecutive MCs or two consecutive OC cycles. This protocol was designed for the IronFEMME project with the aim of stimulating IL-6 production, resulting in the subsequent elevation of hepcidin 3 hours after exercise. ^20^ However, this protocol differs in characteristics with respect to those that have been used to study the bone (re)modelling markers response to exercise, which are typically continuous protocols (60–120 min) and intensity between at 65–75% VO_2max_.^21^

### 2.5. Blood collection

Blood samples were taken between 8 and 10 am to avoid diurnal variability of biochemical parameters.^11^ Intervallic tests were always performed between 8 a.m. and 10 a.m. as well, and the time window was reduced to 1 hour between tests in the different phases of the MC and OC to reduce the intra-participant variability of the results. Two samples (at rest and immediately post-exercise) were drawn from each participant at each MC and OC phase, from an antecubital vein while they were seated to determine the bone (re)modelling marker and sex hormone concentrations. All venous blood samples were obtained using a 21-gauge (0.8 mm × 19 mm, Terumo®) needle. Blood samples for serum variables were collected in a 9 mL Z serum separator clot activator tubes (Vacuette®) and allowed to clot at room temperature for 60 minutes. They were then centrifuged for 10 minutes at 1610 g to obtain the serum (supernatant), divided into 600 μL aliquots, and stored at −80°C.

### 2.6. Blood analysis

17-β-oestradiol, progesterone, P1NP and β-CTX-1 were analysed in serum by electrochemiluminescent immunoassay using Roche Diagnostics reagents in a Cobas e411 Elecsys automated analyser (Roche Diagnostics GmbH, Mannheim, Germany) in the Spanish National Centre of Sport Medicine (Madrid, Spain). Inter-assay and intra-assay CV were: 1.8 and 2.4% at 57.2 ng·ml^-1^ level for P1NP; were 2.1 and 2.8% at 0.403 ng·ml^-1^ level for β-CTX; 11.9% and 8.5% at 93.3 pg·ml^-1^ and 6.8% and 4.7% at 166 pg·ml^- 1^ for 17β-oestradiol; and 23.1% and11.8% at 0.7 ng·ml^-1^ and 5.2% and 2.5% at 9.48 ng·ml^-1^ for progesterone.

### 2.7. Corrections for plasma volume changes

Plasma volume changes (ΔPV) can affect the interpretation of biochemical measurements in blood. In the current study the Dill and Costill equation was used for calculation of the % ΔPV using changes in serum total albumin levels post-exercise in each subject, given their correlation with % ΔPV.^22^ The following equations^22^ for P1NP and β-CTX-1 corrections were used:

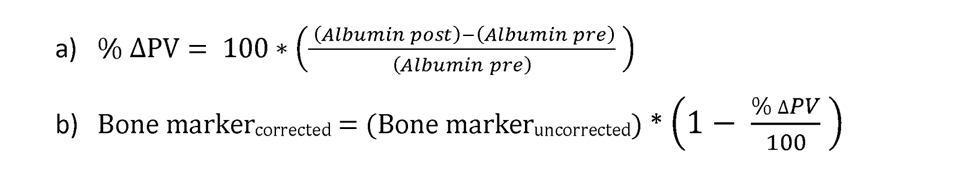

Sex hormone concentrations were not corrected because, although part of the increase in post-exercise circulating hormone concentrations was a result of a decrease in plasma volume, the biological action of these hormones is of greater interest and the concentration of a hormone determines its effect.^23^

### 2.8. Nutritional recommendations

In order to ensure that nutrient intake was not a confounding factor in our results, a nutritionist prescribed the breakfast meal, and participants replicated the same breakfast at least 2h prior to the intervallic tests in all the MC and OC phases before the different blood draws. Nutritional recommendations were standardised 48 h prior and 24 h following the running protocol (for diet composition see Supplementary Material 1).

### 2.9. Statistical analysis

Normality tests were performed using the Shapiro-Wilk test. Data for non-normally distributed variables were log-transformed for analysis.^24^

Participant characteristics were analysed using independent samples t-tests. To explore our objectives, mean concentrations of bone (re)modelling markers and sex hormones were compared between MC phases (EFP vs LFP vs MLP) and OC cycle phases (WP vs APP) using the mixed linear model to analyse repeated measures. The phases and time were set as fixed effects (both intra-subject), and subjects were set as random effect. Comparing hormonal profiles, the mixed linear model analysis was also performed, conducting a separate analysis for each of the following comparisons: EFP vs WP, EFP vs APP, LFP vs WP, LFP vs APP, MLP vs WP, and MLP vs APP. In this case, the ovarian hormonal profile (inter-subject) and time (intra-subject) were set as fixed effects, and subjects were set as random effects. Bonferroni’s post hoc test was applied to pairwise comparisons when the main effect was significant (p<0.05). The effects sizes are reported as partial eta squared (η²p) whose interpretation is 0.01 = small, 0.06 = moderate, 0.14 = large effect. For significant post hoc comparisons Cohen’s d was used and interpreted based upon the following criteria: 0.2 = small, 0.5 = medium, 0.8 = large effect.^25^ Data are presented as mean±1SD.

## 3. Results

### 3.1. Sex hormones

17β-oestradiol showed significant main effects of phase in eumenorrheic females, showing lower 17β-oestradiol levels in the EFP compared to the LFP (p=<0.001; d=-2.099) and MLP (p=0.003; d=-1.731); and time, reflecting an increase from pre-to post exercise. There was, however, no interaction effect (see 17β- oestradiol levels on Table 2). Likewise, progesterone showed a significant main effect of phase, where concentrations were significantly higher in the MLP compared to the EFP (p<0.001; d=-4.047) and LFP (p<0.001; d=-3.381); and time, showing an increase from pre-to post-exercise; but no interaction (see progesterone levels on Table 2).

**Table 2.** P1NP, β-CTX-1, 17-β-oestradiol, and progesterone (mean±SD) of eumenorrheic participants in the different menstrual cycle phases. EFP: early follicular phase; LFP: late follicular phase; MLP: mid-luteal phase. *Significantly different from total EFP. # Significantly different from total LFP. $ Significantly different from total pre-exercise. Significant differences p<0.05.

| *Eumenorrheic females* |  |  |  |  |  |  |  |  |  |
| --- | --- | --- | --- | --- | --- | --- | --- | --- | --- |
|  |  | *EFP* | *LFP* | *MLP* | *Total* | *Main effects* |  |  |  |
|  |  |  |  |  |  |  | *Phase* | *Time* | *Phase\*Time* |
| *P1NP (ng·ml<sup>-1</sup>)* | *Pre* | 55.02±15.76 | 58.87±11.98 | 53.67±11.62 | 55.86±12.86 | **F** | 8.45 | 50.193 | 1.381 |
|  | *Post* | 66.91±16.26 | 80.66±16.35 | 64.57±9.68 | 70.71±15.59<sup>§</sup> | **p** | 0.001 | <0.001 | 0.265 |
|  | *Total* | 60.96±16.64 | 69.97±17.84* | 59.122±11.77<sup>#</sup> |  | **η<sup>2</sup>p** | .414 | 0.939 | .261 |
| *β-CTX-1 (ng·ml<sup>-1</sup>)* | *Pre* | 0.478±0.168 | 0.483±0.197 | 0.388±0.126 | 0.45±0.165 | **F** | 7.929 | .044 | 1.126 |
|  | *Post* | 0.426±0.132 | 0.509±0.140 | 0.364±0.068 | 0.433±0.128 | **p** | 0.001 | 0.835 | 0.336 |
|  | *Total* | 0.452±0.148 | 0.496±0.166 | 0.376±0.098*<sup>#</sup> |  | **η<sup>2</sup>p** | .454 | .007 | .185 |
| 17-β-oestradiol (pg·ml<sup>-1</sup>) | *Pre* | 40.73±33 | 239.94±182.36 | 127.96±45.47 | 136.21±134.23 | **F** | 39.956 | 4.397 | 0.088 |
|  | *Post* | 53.7±46.93 | 369.96±276.18 | 203.159±92.85 | 208.94±209.57<sup>§</sup> | **p** | <0.001 | 0.043 | 0.916 |
|  | *Total* | 47.22±39.75 | 304.95±235.85* | 165.56±80.6* |  | **η<sup>2</sup>p** | 0.702 | 0.924 | 0.209 |
| Progesterone (ng·ml<sup>-1</sup>) | *Pre* | 0.292±0.151 | 1.471±2.845 | 12.463±4.96 | 4.74±6.42 | **F** | 91.145 | 0.783 | 1.404 |
|  | *Post* | 0.749±0.378 | 1.884±2.477 | 13.966±5.109 | 5.533±6.869<sup>§</sup> | **p** | <0.001 | 0.005 | 0.259 |
|  | *Total* | 0.521±0.365 | 1.677±2.586 | 13.214±4.926*<sup>#</sup> |  | **η<sup>2</sup>p** | 0.875 | 0.619 | 0.445 |

On the other hand, in OC users 17β-oestradiol showed a significant main effect of phase, reflecting lower endogenous 17β-oestradiol concentrations in the APP than in the WP (see 17β-oestradiol levels on Table 3); but no main effect of time and no interaction were observed. Endogenous progesterone showed a significant main effect of time, where post-exercise concentrations increased from pre-exercise; but no main effect of phase and no interaction were shown (see progesterone levels on Table 3).

**Table 3.** P1NP, β-CTX-1, 17-β-oestradiol, and progesterone (mean±SD) of oral contraceptive users in the different oral contraceptive cycle phases. WP: withdrawal phase; APP: active pill-taking phase. *Significantly different from total WP. # Significantly different from post-exercise WP. $ Significantly different from total pre-exercise. Significant differences p<0.05.

| *Oral contraceptive users* |  |  |  |  |  |  |  |  |
| --- | --- | --- | --- | --- | --- | --- | --- | --- |
|  |  | *WP* | *APP* | *Total* | *Main effects* |  |  |  |
|  |  |  |  |  |  | Phase | Time | Time*Phase |
| *P1NP (ng·ml<sup>-1</sup>)* | *Pre* | 49.43±6.26 | 47.19±6.1 | 48.31±6.08 | **F** | 45.72 | 14.884 | 26.346 |
| | *Post* | 61.75±8.32 | 45.45±6<sup>#</sup> | 53.6±10.95<sup>\$</sup> | **p** | <0.001 | <0.001 | <0.001 |
|  | *Total* | 55.59±9.54 | 46.32±5.91* |  | **η<sup>2</sup>p** | 0.876 | 0.674 | 0.782 |
| *β-CTX-1 (ng·ml<sup>-1</sup>)* | *Pre* | 0.411±0.136 | 0.368±0.092 | 0.390±0.114 | **F** | 2.516 | 0.353 | 0.106 |
|  | *Post* | 0.415±0.156 | 0.354±0.106 | 0.384±0.132 | **p** | 0.128 | 0.559 | 0.748 |
|  | *Total* | 0.413±0.141 | 0.361±0.096 |  | **η<sup>2</sup>p** | 0.149 | 0.061 | 0.138 |
| *17-β-oestradiol (pg·ml<sup>-1</sup>)* | *Pre* | 32.75±26.47 | 9.60±10.06 | 21.17±22.74 | **F** | 37.899 | 3.148 | 0.058 |
| | *Post* | 60.28±55.06 | 12.16±12.87 | 36.22±45.94<sup>\$</sup> | **p** | <0.001 | 0.091 | 0.380 |
|  | *Total* | 46.51±44.09 | 10.88±11.24* |  | **η<sup>2</sup>p** | 0.653 | 0.875 | 0.674 |
| *Progesterone (ng·ml<sup>-1</sup>)* | *Pre* | 0.304±0.151 | 0.355±0.109 | 0.329±0.13 | **F** | 3.321 | 56.654 | 0.138 |
| | *Post* | 0.772±0.307 | 0.834±0.312 | 0.778±0.305<sup>\$</sup> | **p** | 0.083 | <0.001 | 0.812 |
|  | *Total* | 0.513±0.319 | 0.595±0.335 |  | **η<sup>2</sup>p** | 0.589 | 0.779 | 0.013 |

Comparing sex hormone concentrations between ovarian hormonal profiles, endogenous 17β-oestradiol showed significant main effect of hormonal profile for EFPvsAPP, LFPvsWP, LFPvsAPP, MLPvsWP, and MLPvsAPP analyses, where 17β- oestradiol was higher in these MC phases compared to the APP phases. Moreover, a significant interaction (hormonal profile*time) was observed in LFPvs WP and LFPvsAPP, where pre-and post-exercise 17-β-oestradiol was higher in the LFP compared to WP (pre-: p=0.008, d=1.984; post-: p=0.018, d=1.786) and APP (pre-: p<0.001, d=3.452; post-: p<0.001, d=3.687) (see values in Tables 2 and 3 and statistics in Table 4).

**Table 4.**
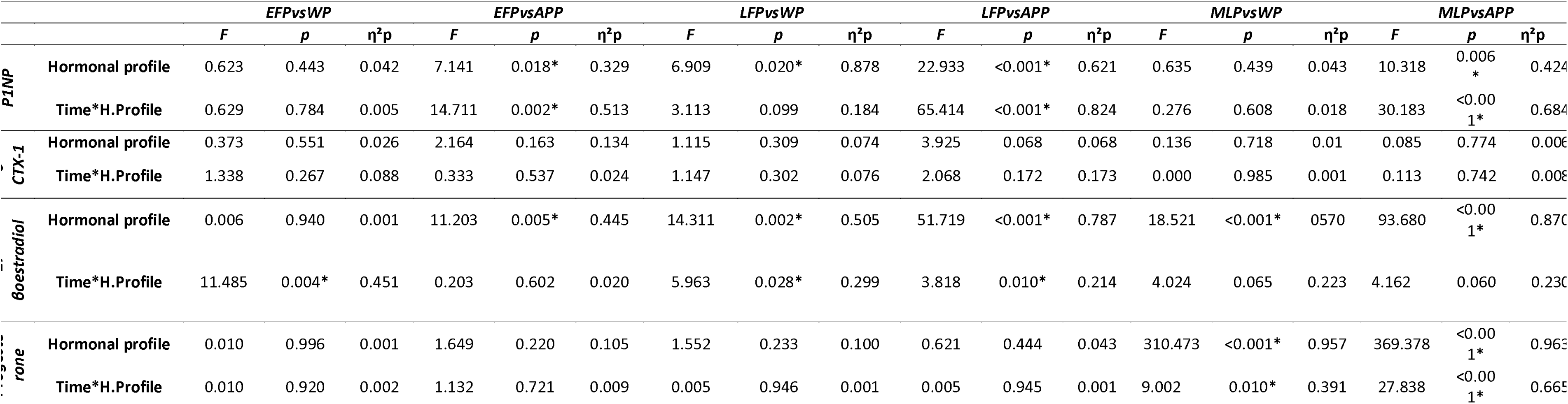
Statistics (F, p and η²p) of each analysis performed to compare the phases of the different ovarian hormonal profiles. EFP: early follicular phase; LFP: late follicular phase; MLP: mid-luteal phase; WP: withdrawal phase; APP: active pill-taking phase. *Significant main effect.

In addition, progesterone showed a significant main effect of hormonal profile for MLPvsWP and MLPvsAPP, where progesterone concentrations were higher in the MLP compared to WP and APP. Furthermore, a significant interaction (hormonal profile*time) was shown for MLPvsWP and MLPvsAPP, where pre-and post-exercise progesterone was higher in the MLP compared to WP (pre-: p<0.001, d=8.144; post-: p<0.001, d=6.467; respectively) and APP (pre-: p=0.001, d=9.888; post-: p<0.001, d=7.871; respectively) (see values in Tables 2 and 3 and statistics in Table 4).

### 3.2. P1NP

In eumenorrheic females, significant main effects of phase and time were observed, but no interaction was shown. Higher P1NP concentrations were shown in the LFP compared to the EFP (p=0.006; d=-0.659) and the MLP (p=0.002; d=0.734). Moreover, post-exercise concentrations were higher than pre-exercise (see P1NP levels on Table 2).

In OC users, significant main effects of phase, showing higher P1NP concentrations in the WP compared to the APP; and time were observed, where post-exercise concentrations were higher than pre-exercise. Moreover, a significant time *phase interaction was shown, highlighting greater post-exercise concentrations in the WP compared to the APP (p<0.001, d=2.419) (see P1NP concentrations on Table 3).

When P1NP levels were compared between participants with different ovarian hormonal profiles (eumenorrheic vs OC users), P1NP main effect of hormonal profile in EFPvsAPP, LFPvsWP, LFPvsAPP, MLPvsAPP was shown; where EFP, LFP and MLP reflected a higher level of P1NP compared to APP, while LFP showed a higher concentration in comparison with WP. In addition, a significant hormonal profile*time interaction was shown in EFPvsAPP, LFPvsAPP and MLPvsAPP; showing lower post-exercise P1NP concentrations in APP compared with EFP (p<0.001; d=1.879), LFP (p<0.001; d=3.371) and MLP (p=0.002; d=2.311), without significant differences between pre-exercise P1NP concentrations between MC phases and APP (see concentrations in Tables 2 and 3 and statistics in Table 4).

### 3.3. β-CTX-1

There was a significant main effect of phase for β-CTX-1 in eumenorrheic females, where pairwise comparisons reflected lower concentrations in the MLP compared to the LFP (p=0.001; d=0.804) and the EFP (p=0.039; d=0.540). No other main effects or interactions were shown (see values in Tables 2 and 3 and statistics in Table 4).

## 4. Discussion

This study investigated β-CTX-1 and P1NP responses to intervallic running throughout the MC and OC phases, while comparing ovarian hormonal profiles. Bone formation, measured by P1NP concentrations, and bone resorption, determined by β- CTX-1, were affected by MC-related fluctuations following exercise, showing greater bone formation in the LFP and reduced bone resorption during the MLP. These results are not in line with those shown by Guzman et al.^12^ where no differences were shown between MC phases [mid-late follicular phase (day 8±1) and luteal phase (day 22±3)] in a group of seven eumenorrheic females. Nevertheless, it should be noted that the Guzman et al.^12^ study did not measure the EFP (in this study day 4±1) nor the LFP (as defined in the present study as 1-3 days before the ovulation day, day 12±2), but rather measured the mid or late follicular phase, depends on the individual characteristics of each female’s MC. Moreover, luteal phase testing was scheduled 1 week after a positive LH detection kit test (day 22±3) in the Guzman et al.^12^ trial, coinciding with our timing of MLP measurement (day 21±3). These differences in the timing of measurement during the MC between the present study and the Guzman et al.^12^ trial could explain the discordant results, since the participants included in the Guzman et al.^12^ study were not evaluated when progesterone concentrations were as low as our participants in the EFP (follicular phase^12^: 4.12±2.36 ng·ml^−1^) and did not reach as high 17β-oestradiol values in the follicular phase^12^ (46.3±8.9 pg·ml^−1^) or the luteal phase^12^ (67.3±23.4 pg·ml^−1^) as our participants in the LFP (see Table 2). Therefore, the fact that the present study showed a significant main effect of MC phase in P1NP in contrast to Guzman et al. may be supported by the known beneficial effect of 17β-oestradiol on bone metabolism in addition to the recognized role of progesterone as an 17β-oestradiol partner in bone metabolism, which has been shown to promote bone formation by increasing the number, maturation, and differentiation of osteoblasts *in vitro*.^26^

Regarding results from OC users, no differences in P1NP and β-CTX-1 levels were observed at rest between OC phases (see days on Figure 1), in contrast to the results shown by He et al.^27^ in which β-CTX-1 concentrations were lower in the mid APP (day 22 to 28) and P1NP concentrations were lower in the mid and late APP (day 10 to 26) at rest. Moreover, the results presented herein disagree with the Martin et al.^13^ study, in which β-CTX-1 were lower in the APP (days 15-16) compared to the WP (days 3-4) at rest. It is worth noting that the participants included in the He et al. and Martin et al.^13^ studies used a specific formulation of OCs containing 30 μg ethinyl oestradiol and 150 μg levonorgestrel, as opposed to the participants included in the present study who did not standardize the composition and dosage of OC they used, which may explain the difference in resting results.

Additionally, this study is the first to show bone (re)modelling marker concentrations after exercise in OC users. The main finding is the greater increase in post-exercise P1NP in the WP compared to the APP following the same protocol, which may suggest the same exercise protocol may involve a different stimulus between phases of the OC cycle. This non-variation of post-exercise P1NP in APP, when 17β- oestradiol concentration is lower, could be linked to the existence of an 17β-oestradiol concentration threshold, which other studies hypothesize to be approximately 26-31 pg·ml^-1^, representing the threshold level below which oestrogen receptor α (ERα) in bone cells are not occupied by oestrogens, leading to a skeletal functional oestrogen deficiency.^28^ Thus, the findings from the present study appear to support other studies conducted in *in vitro* models showing that the ERα is involved in the osteogenic response to mechanical stress, thus low concentrations of 17β-oestradiol could reduce the mechanosensitivity of osteocytes and the responsiveness of bone cells to mechanical load.^29^ Nonetheless, exogenous sex hormones must be taken into consideration. Although ethinyl oestradiol shows a similar affinity to 17β-oestradiol to ERα,^30^ other factors could intervene in its effect on bone metabolism such as the low dose of ethinyl oestradiol contained in these OCs, the possible binding of progestins to the ERα^31^ and the increase that other studies reported in levels of sex hormone-binding globulin,^32^ decreasing the bioavailability of ethinyl oestradiol to bind to the ERα. Therefore, other factors associated with these synthetic hormones could mediate bone metabolism, decreasing the P1NP response to this running protocol.

Comparing bone (re)modelling markers between participants with different ovarian hormonal profiles, lower post-exercise P1NP concentrations were shown in the APP of the OC cycle compared to the EFP, LFP and MLP of the MC without significant differences in pre-exercise P1NP levels. Furthermore, significant differences between pre-and post-exercise in P1NP concentrations in all MC phases versus no difference between pre-and post-exercise in the APP of the OC cycle can be observed. Thus, considering the existence of significant differences between pre-and post-exercise values of 17β-oestradiol in the MC phases cycle versus the absence of differences in pre-and post-exercise values in OC users, it could be suggested that the increase in post-exercise P1NP concentrations could be associated with this increase in 17β - oestradiol, apart from the stimulus derived from exercise. This fact reinforces the role of ovarian sex hormones, especially 17-β-oestradiol, in the osteogenic response to an exercise stimulus.^28,29,33^ In addition a significant main effect has also been shown, where higher P1NP levels were observed in the LFP of the MC compared to the WP of the OC cycle. Thus, the higher P1NP levels observed in the LFP could be positively influenced by the higher 17β-oestradiol concentrations, which may contribute to higher bone formation.^28,29,33^ This higher bone formation might not be achieved by OC users due to its lower levels of 17β-oestradiol. These findings may provide some evidence of differences in bone metabolism in females with different ovarian hormonal profiles. Nonetheless, these results need to be supported by long-term studies conducted with healthy OC users using additional methods of bone health assessment (*i.e.* DXA or Quantitative Computed Tomography) to examine the possible effects that this difference in acute exercise response may have on bone health in the long-term, since an association has been observed between exposure to the use of hormonal contraceptives and BMD, which should be taken into account when assessing the OC effect on bone health.^34,35^

P1NP concentrations increased post-exercise, whereas β-CTX-1 values did not vary significantly in response to exercise in both groups. Although the running protocol performed in this study (intervallic at 85% VO_2max_) differs from others reported in the literature (continuous protocols between 60-120 min and intensity at 65-75% VO_2max_)^21^, the increase in P1NP appears to be in line with other studies in which participants have performed running protocols.^12,36^ While some investigations have reported post-exercise β-CTX-1 data in which the concentration did not vary,^37^ agreeing with the present results, others have shown a decrease in this bone resorption marker.^12^ Despite the fact that P1NP is considered an indicator of bone formation, some authors have suggested that transient increases in P1NP may be related to exercise-induced damage resulting in a small release of connective tissue content into the blood.^21^ Nevertheless, controversy still surrounds the microdamage repair mechanism suggesting that there are alternative mechanisms of direct repair of the bone matrix that dońt need to involve removal and replacement of bone by remodelling.^38^ In fact the results of Seref-Ferlengez et al.,^38^ suggest that alternative repair mechanisms exist in bone to address matrix micro-cracks *in vitro*, given that previously damaged bone tissue recovered control values 14 days after damage occurred. This alternative mechanism may also explain this increase in post-exercise P1NP without any variation in β-CTX-1. Although the results of the present study show an increase in bone formation after exercise, this fact may not imply a long-term osteogenic effect, as there is still limited evidence to interpret these bone (re)modelling marker data and some studies suggest that endurance running training may decrease spine BMD.^34,39^ Therefore, a long-term follow-up should be performed to really draw conclusions about the stimulus of running training on bone health and to establish a relationship with the increase in P1NP after high-intensity exercise.

The main strength of this study is the consideration of the hormonal environments throughout the MC and OC cycle, by measuring serum 17β-oestradiol and progesterone, and using ovulation tests to measure LH surge according to guidelines.^19^ In addition, exercise trials were performed in the morning, with a maximum interval of one hour between trials, using standardized protocols and indications^11^ for the preservation and measurement of serum sex hormones and bone (re)modelling markers to avoid within-and between-subject variability. Furthermore, this original research could expand knowledge on this topic, as a recent systematic review with meta-analysis only included studies on P1NP and/or β-CTX-1 responses to running exercise in healthy young adult males, evidencing the lack of similar studies in female populations, especially in premenopausal females.^21^ Nevertheless, there were some limitations, such as the fact that a specific type of OCs with standardized composition and doses of synthetic hormones was not used. Given the different properties of different synthetic progestins in terms of binding affinities and transcriptional activities when binding to androgen or oestrogen receptors, there could be a different magnitude of effect and biological consequence.^31^ Finally, it should be mentioned that although endogenous sex hormones have been measured in serum and in the case of OC users the OC dosages have been reported, a good practice could be to measure the synthetic sex hormone concentrations in serum.

## 5. Conclusions

MC phase affected bone (re)modelling markers by showing higher bone formation, measured by P1NP concentrations, in the LFP and lower bone resorption, as measured by β-CTX-1, during the MLP. OC users showed decreased P1NP levels post-exercise in the APP without differences in pre-exercise levels, when endogenous 17β- oestradiol was lower and exogenous ethinyl oestradiol and progestins were higher. Moreover, a different behaviour of P1NP in post-exercise was seen between eumenorrheic females in all MC phases, where a significant increase in P1NP was shown, and OC users in the APP, where no post-exercise increase was observed. These findings underscore the importance of studying exercising females with different ovarian hormone profiles, as these changes in sex hormone concentrations affect bone metabolism in response to high intensity running exercise and could have long-term implications for bone health that should be studied. Therefore, since exercise is one of the stimuli that can influence bone health in female athletes, and as observed in this study, different sex hormone concentrations influence the acute response to a running stimulus, these two groups of female athletes should be studied independently if the objective is to assess bone health.

## Data Availability

The data that support the findings of this study are available from the corresponding author, upon reasonable request

