## Supplementary Material 1 for "Influence of menstrual cycle and oral contraceptive phases on bone (re)modelling markers in response to intervallic running"

**Nutritional recommendations**

Breakfast compositions were scrambled eggs with spinach and a piece of fruit or three pieces of fruit with 50 g of dried fruit (e.g., walnuts) and wholegrain (>70% of the product must have whole grains) toast with one teaspoon of tahini or peanut butter. In addition to the standardized breakfast, some recommendations were provided for the other meals, as follows: lunch/dinner: 100% whole grain cereals (*e.g.*, barley) or potato/sweat potato baked, or legumes, with poultry, plant-based protein (*e.g.*, tofu), fish or seafood, with a mix of a cooked and raw vegetables; Cooking oils were limited to extra virgin olive oil or extra virgin avocado oil; Beverages: water; Snacks: pistachios with citrus fruit (no juices were allowed) or a tuna sandwich with wholegrain bread or raisins and nuts.
